# Assessing the Perception of Usefulness and Use of Syndromic Surveillance of Surgical Site Infections for Improvement of Infection Prevention and Control in Health Facilities of Tanzania: A Qualitative Design’ for Publication

**DOI:** 10.64898/2026.11.16.26368239

**Authors:** Joseph C. Hokororo

**Affiliations:** Health Quality Assurance Unit, Ministry of Health, Government City – Mtumba, Afya Road, P.O.BOX 743, 40478, DODOMA - Tanzania; University of Central Nicaragua, Campus Central, Semoros del Zumen, 3 C. Abajo, 1 C. al Lago, Telonos: 2279-1160 / 2269-3103, Nicaragua; Texila International Marketing Management Academic Delivery Facilitator of Central University of Nicaragua, Office 1 & 3, 5th Floor, ELCOTSEZ, ELCOT IT Tower, Vilankurichi, Aerodrome Post, Coimbatore-641014, Tamil Nadu, India

## Abstract

Tanzania has recently set up a syndromic surveillance of Surgical Site Infections (SSIs) system with the aim to determine the burden of SSI in the country. This study Assessed the Use and Usefulness of it for Infection Prevention and Control (IPC) improvement in health facilities of Tanzania. The study utilized the classic Technology Acceptance Model (TAM) adapted specifically for healthcare information systems; the research team developed semi-structured Key Informants Interview (KII) guides. The QIT members were purposefully selected as KIIs in PHFs. Data collection continued until saturation was achieved, ultimately gathering insights from a total of 49 participants across 20 PHFs in Tanzania from September to November 2025. The interviews were audio-recorded and lasted for around 60 minutes. Privacy was ensured by conducting the interview in an isolated room. Textual data were coded and categorized using NVivo version 20.2 (NVivo.exe). The QIT members reported their perception that syndromic surveillance helped to track SSIs, helped to get burden and trends, and were useful for planning. Participants generally liked the syndromic surveillance and reported using them to inform program design, guide development of intervention and enhance personal knowledge. The participants shared important feedback to improve the syndromic surveillance, particularly as a need to enhance improvement of IPC. Tanzania’s syndromic surveillance system use for tracking SSIs in primary care is recognized by QIT as a transformative tool for mapping SSI burdens and informing strategic planning. To ensure long term success, stakeholders must address feedback by strengthening quality of data, providing continuous training, and adapting the system to meet local needs. The overall results of this study showed the importance of conducting syndromic surveillance in order to understand the burden of SSI so as to come up with mitigation strategies of improving IPC across the health system.

## 1. INTRODUCTION

The burden of healthcare associated infections (HAIs) remains a critical threat to global patient safety, with the majority coming from low- and middle-income countries (LMICs). Among these, SSIs are the most frequent reported HAI in resource constrained surgical settings, directly increasing patient morbidity, hospital stay duration, financial strain, and mortality(1,2). In Sub-Saharan Africa, where surgical bottlenecks are immense and perioperative resources are strained, SSI rates soar far above global averages, frequently ranging between 10% and 24% in tertiary and regional referral facilities, that means at Primary health facilities (PHFs) the situation might be even worse(3–5). This high disease burden reflects an urgent need for robust infection prevention and control (IPC) frameworks(6). However, the foundational pillar of any IPC framework systematic, data driven disease surveillance is routinely crippled in these environments by systemic infrastructural limitations.

Traditional microbiological surveillance represents the gold standard for tracking HAIs, yet its execution requires capital intensive laboratory infrastructure, consistent supplies of reagents, and specialized personnel(7). In public health facilities across Tanzania, these requirements are rarely met sustainably. Clinical decisions and infection tracking are routinely forced to bypass laboratory confirmation due to frequent stockouts, inadequate training, or prohibitive costs passed down to patients. Consequently, relying strictly on culture-based confirmation results in a severe underreporting of SSIs. This data void prevents facility management, IPC committees, and national policymakers from understanding the true magnitude of the crisis, identifying specific problem wards, or evaluating the efficacy of existing hygiene protocols. Without reliable, timely data, IPC improvement initiatives remain reactive, uncoordinated, and largely ineffective.

To bridge this critical infrastructure gap, syndromic surveillance has emerged as a pragmatic and highly valuable strategy. Tanzania has introduced the syndromic surveillance for SSI because of limited culture and sensitivity infrastructure. This methodology shifts the focus from definitive laboratory pathogen identification to the systematic, standardized tracking of clinical signs and symptoms such as localized pain, purulent drainage, erythema, and localized swelling. By utilizing standardized clinical definitions that frontline health workers can apply at the bedside, syndromic surveillance bypasses laboratory bottlenecks. It offers a low cost, real-time alternative capable of generating immediate actionable data. In Tanzania, efforts to digitize health information have successfully integrated various disease tracking parameters into national platform, the District Health Information Software 2 (DHIS2). However, while the technical capacity to collect syndromic SSI data exists, the actual deployment, local ownership, and clinical utility of this data within health facilities remain poorly understood.

The core challenge of syndromic surveillance lies not in data collection, but in data use and usefulness perception of the users. For data to drive IPC improvements, healthcare providers must perceive the information as accurate, actionable, and directly beneficial to them and patient care. Too often, electronic health systems degenerate into burdensome, one-way reporting pipelines where overworked clinicians log data for national aggregates without receiving local feedback. If frontline nurses and surgeons view syndromic surveillance forms as an administrative duty rather than a tool for clinical decision making, data quality degrades. This leads to under reporting, misclassification, and missed opportunities for early clinical intervention. Understanding how Tanzanian health workers interact with, interpret, and leverage symptom-based data is essential to transforming passive reporting into active IPC.

This study employs a qualitative research design to examine the perception and organizational factors governing the implementation of syndromic SSI surveillance in Tanzanian health facilities. Quantitative metrics can chart the rise and fall of infection rates, but they fail to capture *why* data is ignored or *how* a clinical team successfully translates a spike in symptom reports into a modified ward cleaning routine. By gathering rich narrative insights from surgeons, nurses, IPC coordinators, and QIT, this qualitative approach explores the lived experiences of use and usefulness of syndromic surveillance of SSI. It uncovers the cultural dynamics of hospital hierarchies, the cognitive workloads of Health care workers (HCWs), and the institutional constraints that dictate whether surveillance data translates its use and usefulness into improved patient safety.

The study will utilize the classic Technology Acceptance Model (TAM) adapted specifically for healthcare information systems. TAM provides a validated structural framework to assess how healthcare personnel perceive the implementation of syndromic surveillance for SSIs(8,9). By isolating two core determinants perceived usefulness (PU) and perceived ease of use (PEOU), the model explains the behavioral intention to adopt this tool for IPC in Tanzanian PHFs. Given the study’s qualitative design, TAM will not be used for statistical testing, but rather as a thematic guide to explore the specific user experiences influencing actual system adoption(10,11)

Ultimately, this investigation seeks to provide a comprehensive, contextualized evaluation of the use and practical usefulness of syndromic surveillance for SSIs in improving IPC. By documenting the use and usefulness of symptom-based tracking within the Tanzanian context, the findings will offer actionable insights for hospital administrators, national policymakers, and international health partners. Strengthening the feedback loop between data collection and clinical practice is vital to reducing the preventable burden of SSIs. This research builds a necessary evidence base to transform routine surveillance from a static bureaucratic exercise into a dynamic, lifesaving engine for continuous IPC improvement. Therefore, this study Assesses the perception of QIT members and HCWs at large on Use and Usefulness of Syndromic Surveillance of SSI for IPC Improvement in Health Facilities of Tanzania.

## Background of Technology Acceptance Model

The TAM defines how individuals adopt or reject information technology based on two primary cognitive beliefs: PU, the degree to which a user believes a system will enhance their job performance, and PEOU, the extent to which they expect the system to be free of effort. While originally engineered by Fred Davis in 1989 as a quantitative instrument reliant on structured psychometric survey scales, TAM is widely applied in qualitative research as a deductive conceptual lens (10,12). Instead of calculating statistical correlations between variables, qualitative investigators use TAM’s core principles to build semi-structured interview guides and focus group protocols, translating rigid constructs into open-ended questions that explore user attitudes, frustrations, and behavioral intentions regarding a new digital intervention. During data analysis, TAM functions as a structured coding framework for directed content analysis, allowing researchers to categorize transcript data into distinct themes representing PU (such as time-saving features or improved clinical communication) and PEOU (such as interface confusion or intuitive navigation) (13). Furthermore, a qualitative approach to the TAM allows researchers to diagnose the specific, real-world factors that shape user perceptions. It reveals how systemic issues like inadequate workplace training or low digital literacy directly damage Perceived Use and how tools that disrupt daily clinical workflows fundamentally undermine Perceived Usefulness. Rather than relying on rigid adoption data, gathering narrative feedback exposes the psychological and operational mechanics. This deeper insight helps uncover critical external influences including trust, data privacy, and workplace culture enabling researchers to refine and scale the TAM framework to fit highly complex medical environments. The summary of TAM is shown in figure 1.

**Figure 1:**
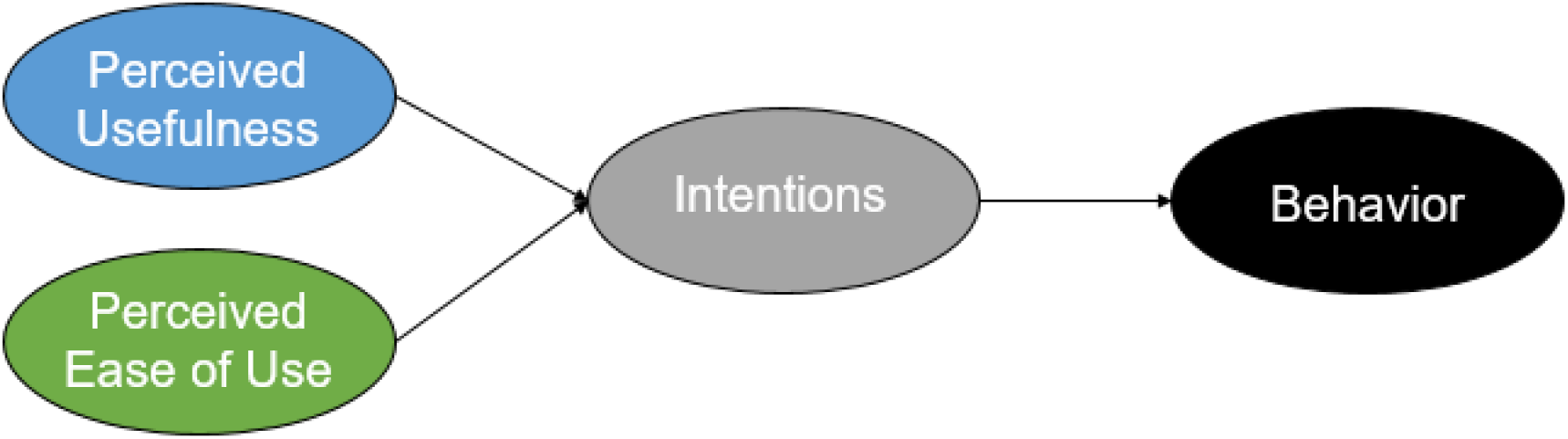
The Technology Acceptance Model (14)

## Materials and Methods

### Study Design

This study employed a qualitative descriptive design utilizing the Technology Acceptance Model (TAM) as an analytical framework. This approach allowed for an in-depth exploration of QIT members lived experiences, perceptions, and attitudes toward the implementation of syndromic surveillance for SSI in Tanzanian PHFs. By using a semi-structured interview guide, we aimed to explore the perceived usefulness and use of this syndromic surveillance system for IPC improvement across health facilities in Tanzania.

### Study Setting

The study was conducted at PHFs of Tanzania which is in the Eastern part of Africa. Tanzania has a total area of 945,087 sq.km including 61,000 sq. km of inland water. Tanzania’s approximate total population was 67,438,106 million as of January 2023. Tanzania has 26 Regions, 139 districts, 184 councils and 12,319 villages and more than 11,000 health facilities (15). Tanzania’s Primary Healthcare (PHC) framework functions as a decentralized pyramid from community to district hospitals. The health system of Tanzania had been divided into primary, secondary and tertiary levels of healthcare(16). This design provides standardized packages of preventative and curative services at each tier.

Administrative oversight for PHC rests with the Prime Minister’s Office for Regional Administration and Local Government through District Medical Officers. Levels of PHC are as follows: Community Level - Community health workers manage community health promotion, education, and early triaging; Dispensary Tier - Serves as the primary village entry point. This tier treats a population of 6,000 to 10,000 people exclusively via outpatient basic care; Health Centers - operates at the ward level to serve roughly 50,000 people. This tier offers 24/7 inpatient services and critical maternal care; Council Hospitals - acts as the highest primary tier authority. It handles advanced diagnostic care and surgical operations before cascading patients to regional hospitals.

### Study Population

Our study population were all QITs who are involved in surgical care safety and IPC in PHFs who were at the facility during the days of data collection in Tanzania.

### Selection Criteria

#### Inclusion

The PHFs that have operating theaters and conducted surgery in the past three months at the time of data collection. All QITs in PHFs who were at work during the days of data collection and provided informed consent in Tanzania.

#### Exclusion Criteria

All QITs in PHFs who were not at the facility during the days of data collection.

#### Sampling technique

This study used PHC of health care provision to study the Use and Usefulness of Syndromic Surveillance of SSI for IPC improvement in Tanzania from the QIT members. A multistage cluster sampling procedure was used to select representative facilities from nine zones of Tanzania. However, the standard of healthcare provision was more or less homogeneous across the zones.

The country was divided into nine administrative zones involved in sampling: 1. central zone with 3 regions (Dodoma, Singida and Manyara), 2. Special Dar es Salaam Zone with 3 health sub-regions (Ilala, Temeke and Kinondoni), 3. Eastern zone with two regions (Morogoro and Pwani), 4. Lake zone which had six regions (Geita, Kagera, Mara, Mwanza, Shinyanga, and Simiyu), 5.

Northen zone which had three regions (Arusha, Kilimanjaro, and Tanga), 6. Southen zone had three zones (Lindi, Mtwara and Ruvuma), 7. Southern highlands zones which had two regions (Iringa and Njombe), 8. South West highlands with two regions (Mbeya and Songwe) and 9. Western zone which had three regions (Rukwa, Katavi, Tabora and kigoma).

The first sampling stage involved creating a sampling frame of zones. In each zone one region was picked while in lake zone which has many more regions than others, two regions were picked that and therefore, 10 regions out of 26 regions were sampled. The simple random sampling was done by lottery method to pick regions names from each zone one by one.

The second sampling stage involved PHFs selection from the selected regions. The primary level health facilities sampling frames were created using district hospitals for each region and from which two (2) district hospitals in each sampled region were picked. Two district hospitals were randomly selected from each participating region using lottery method. Hence in total there were 20 primary health facilities two per each region were picked.

The QIT members were purposively selected from the sampled PHFs. We selected all QIT who were at work during the days of data collection. The QITs served as Key Informants (KI) during the interview to assess their understanding on the Use and Usefulness of Syndromic Surveillance of SSI for IPC improvement in health facilities of Tanzania.

### Sampling

We determined by data saturation, typically it happened at15th to 20th participant in every guiding question across the selected PHFs.

### Data Collection

We conducted Key Informant Interview (KII) with the QIT members of the selected PHFs after obtaining the informed consent from them. The KII was guided by using the Semi-structured interview guide developed based on core TAM constructs Perceived Usefulness and Perceived Use. The interviews were audio-recorded and lasted for around 60 minutes. Privacy was ensured by conducting the interview in an isolated room without the presence of other people. Participants were ensured of the confidentiality of the information obtained through the interview. At the end of the interview, a summary was presented to the participants for validation.

### Data Analysis

We used Framework analysis method guided by the pre-defined TAM thematic framework. Interviews were transcribed verbatim within the same day of the interview to prevent the loss of information. Transcripts were reviewed by another person to decrease the bias and increase the interpretive credibility. NVivo version 20.2 (NVivo.exe) was used to derive in vivo codes organized and analyzed text and audio data through the process of qualitative coding using both inductive (open coding) deductive coding (based on themes previously identified during the interview guideline design stage and review of relevant literature). Thematic saturation was defined as when new received information produced little to no new information on the topic under exploration (17). The average saturation was reached when conducted 15th interview. The codes were used to develop categories which eventually were used to come up with thematic content which derived the themes and subthemes as shown in table 1 below. The findings were reported by using consolidated criteria for reporting qualitative research (18). The recorder for the qualitative data was well stored. Descriptive quotations were used in the results part to represent the relevant themes, with perspectives clearly marked. However, improve clear legibility, minor grammatical alterations were applied to some quotes.

**Table 1:**
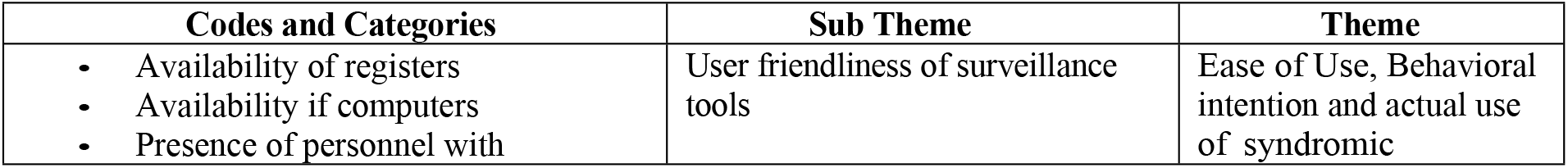
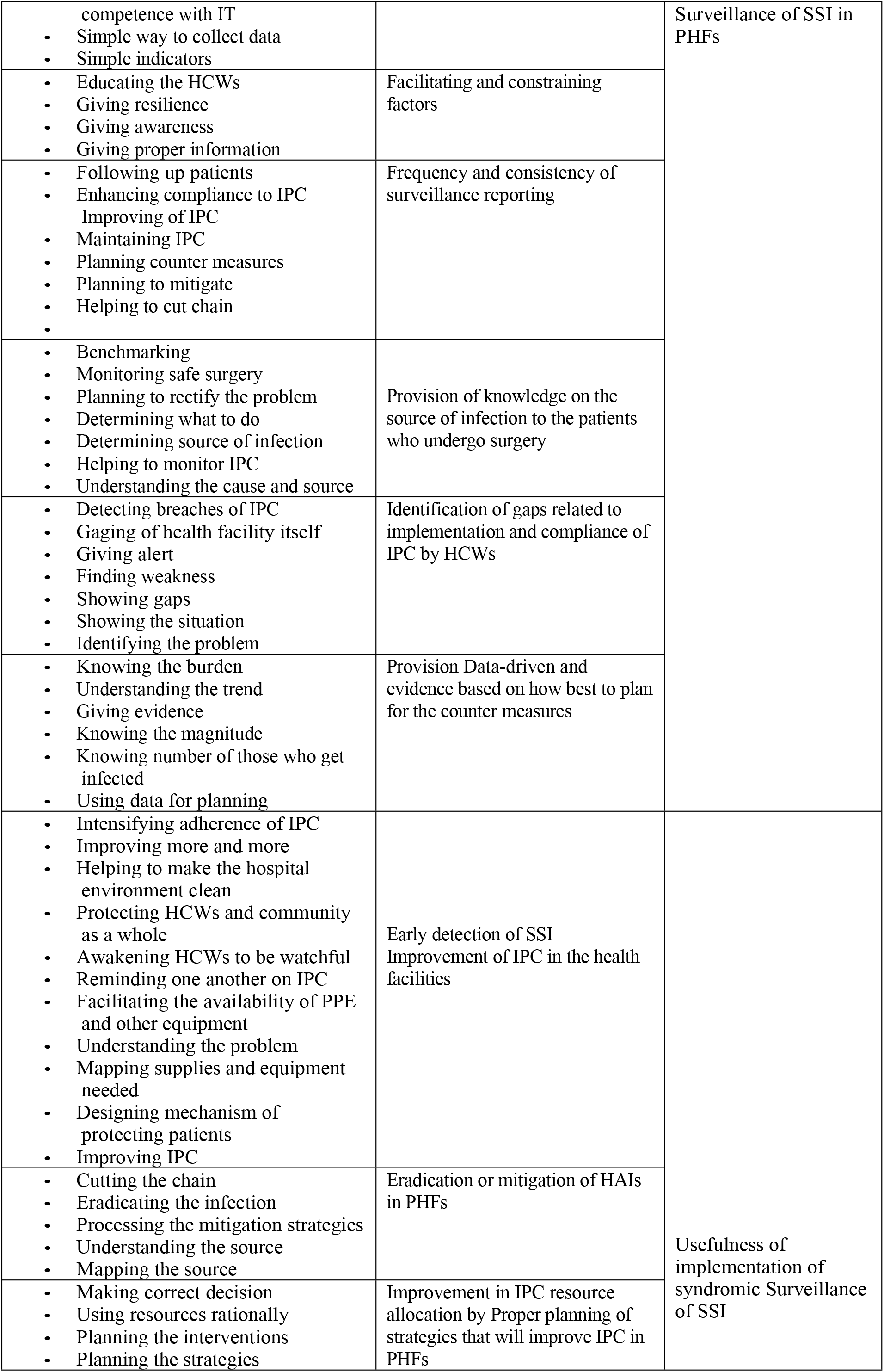
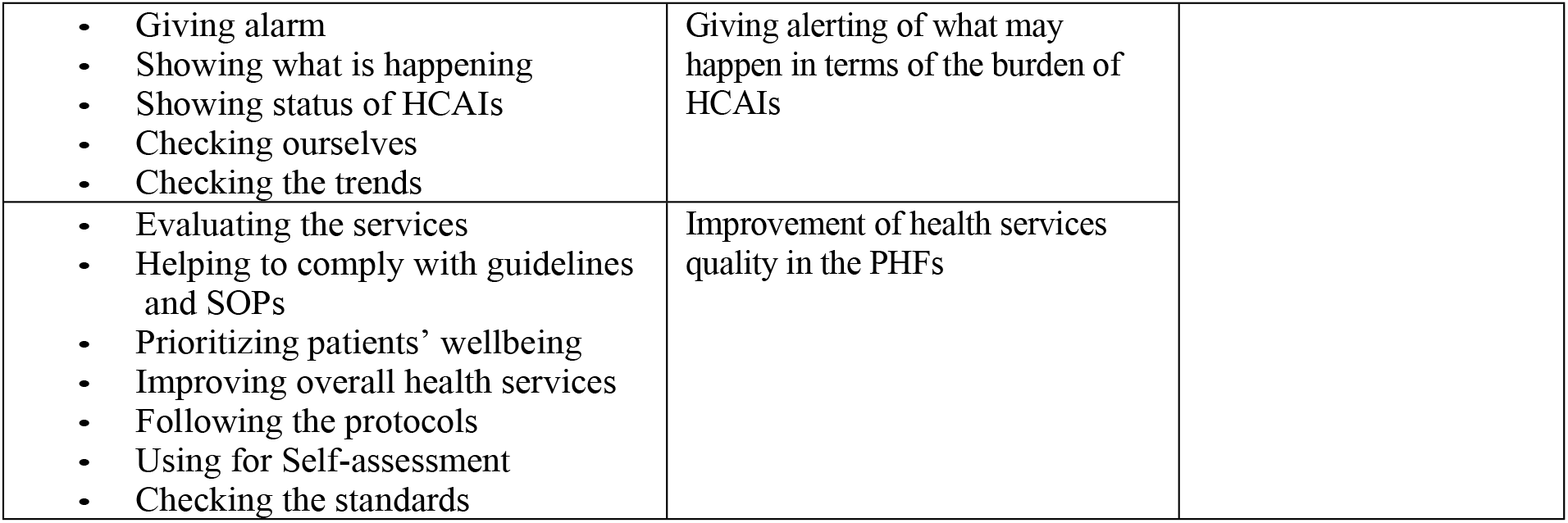
Codes, Categories, Sub Themes and Themes Developed.

### Emergent Themes

This study generated four themes, namely: Use of syndromic Surveillance of SSI in PHFs, Usefulness of implementation of syndromic Surveillance of SSI, Details on how the themes were developed is stipulated in Table 1 below.

## Results

### Characteristics of Study Participants

A total of 49 QIT members from 20 primary healthcare facilities across 10 regions participated in the study. Participants were predominantly nurses (65.3%), followed by medical doctors (18.4%), laboratory technologists, environmental health officers, nutritionists, physiotherapists, social welfare officers, and hospital secretaries. Most participants were female (57.1%), aged between 30 and 59 years (89.8%), and had more than five years of professional experience (77.6%), reflecting substantial experience in quality improvement and infection prevention and control activities.

Participant characteristics are presented in Table 2.

**Table 2.** Socio-demographic characteristics of study participants (N=49).

| **Variables** | **Attribute** | **Number (%)** |
|---|---|---|
| **Age (years)** | 18-29 | 05 (10.2) |
|  | 30-59 | 44 (89.8) |
| **Sex** |  |  |
|  | Female | 28 (57.1) |
|  | Male | 21 (42.9) |
| **Education level** |  |  |
|  | Degree and above | 18 (36.7) |
|  | Diploma and below | 31 (63.3) |
| **Profession** |  |  |
|  | Nurse | 32 (65.3) |
|  | Medical doctor | 09 (18.4) |
|  | Laboratory Technologist | 02 (04.1) |
|  | Environmental officer | 02 (04.1) |
|  | Social well fare Officer | 01 (02.0) |
|  | Nutritionist | 01 (02.0) |
|  | Physiotherapist | 01 (02.0) |
|  | Hospital Secretary | 01 (02.0) |
| **Experience (years)** | 5 year and above | 38 (77.6) |
|  | Less than 5 years | 11 (22.4) |

### Structural framework for TAM results

This study utilised the qualitative approach exploring how participants experienced and perceived the adoption of the syndromic surveillance, grounded in the core conceptual dimensions of the TAM. The empirical data gathered from KII and thematic analysis revealed subjective insights into user motivations, anxieties, and behavioral drivers. The findings were structured around emergent themes that correspond to perceived use, perceived usefulness, and the emotional and contextual factors shaping users’ overall attitudes and behavioral intentions. By capturing the authentic voices and lived experiences of the participants, this analysis provides a deep contextual understanding of the specific factors that influence real-world technology utilization.

The primary purpose of SSI syndromic surveillance within IPC is to track and report infections that arise following surgery. Ultimately, the data was used to prevent SSIs and enhance patient outcomes and safety by comparing infection rates and pinpointing areas that need improvement. The two (2) core themes were abstracted from eleven (11) sub-themes as stipulated in Table 1.

### First Theme: Perceived Usefulness (PU) of implementation of syndromic Surveillance of SSI in PHFs

This theme was conceptualized from five (5) sub-themes, namely Improvement of IPC in the health facilities, Eradication or mitigation of HAIs in PHFs, Proper planning of strategies that will improve IPC in PHFs, giving alert of what may happen in terms of the burden of HCAIs and Improvement of health services quality in the PHFs. The main usefulness of syndromic SSI surveillance is detecting and treating immediately because it can serve as an early warning system for possible outbreaks. This early discovery enables the quick application of infection control strategies, which may stop the infection from spreading further and lessen patient damage. This section reports how QIT members perceive the syndromic surveillance system’s impact on their clinical efficiency of surgical patients and IPC outcomes.

### Subtheme A: Early detection of SSI is useful in Improvement of IPC in the health facilities

This study found syndromic surveillance system was useful for HCWs to identify potential infections faster than traditional laboratory methods. Improving IPC in HFs is greatly aided by SSI surveillance. One participant said “…..*By gathering and examining data on surgical site infections early, it becomes easier to pinpoint areas that need improvement fast, enables focused treatments, and eventually lowers the frequency of these infections and hence, improving patient outcomes*….QIT30” Another of the participants highlighted that… “*surveillance is useful because instead of putting broad, sometimes ineffectual measures into place, healthcare facilities may concentrate their resources and efforts on the areas that most require improvement and hence Intensifying adherence of IPC and improve more and more….* QIT01*”* Other participants said…. “*SSI surveillance’s usefulness is on prioritization of hospital cleanliness, especially in the operating room and wards…* QIT02*”*. That is what one of the participants emphasized…. “*With an emphasis on keeping the hospital environment clean, successful SSI surveillance necessitates monitoring infections, determining risk factors, and assessing the success of preventative actions*…. QIT03”.

SSI surveillance is useful for safeguarding healthcare workers (HCWs) and the general public when the pathogens are identified early. The respondents felt that…. “*Surveillance is useful in reducing the possibility that patients will transfer pathogens into the community when early identified and removed*…. QIT04” Also, SSI surveillance is useful for making HCWs more vigilant. By giving HCWs the information and insights they need to monitor infection rates, pinpoint risk factors early, and put focused preventative measures into place. Another respondent said…. “*By conducting SSI surveillance, HCWs can be more watchful in their procedures, which will ultimately identify SSI early and measures are taken early as well*…. QIT05” The surveillance is useful for reminding HCWs of best practices, assess the success of implemented measures, and identify areas where IPC practices need to be strengthened by tracking SSI data. One of the respondents said…. “*The initiative is useful because it determines risk factors and put specific preventative measures into action*…. QIT3”. In the end, this proactive strategy lowers the incidence of SSIs by empowering healthcare providers by reminding one another on adherence of IPC measures.

### Subtheme B: Eradication or mitigation of HAIs in PHFs

Surveillance is useful part of multifaceted strategy centered on prevention and control required to eradicate or mitigate the HAIs through adopting suitable IPC procedures, encouraging antimicrobial stewardship, keeping a clean and hygienic atmosphere, and practicing strict hand hygiene are important tactics. One member of QIT said…. “*You have to know the burden of the problem like SSI magnitude for you to make decision on how to mitigate or eradicate* …. QIT06” Another participant said…… “*In the context of SSI surveillance, it is useful for breaking the infection chain, stopping the transmission of SSIs, and eventually mitigating their incidence. This entails carrying out a number of interventions and monitoring initiatives in order to detect, manage, and stop SSIs*…. QIT07”

In order to define and improve mitigation methods, SSI surveillance is useful for determining infection rates, benchmarking, initiating clinical evaluations, and putting infection control measures into place. One of the QIT members agreed that… “*Healthcare facilities is useful in identifying problem areas and assess the success of applied mitigation strategies by monitoring SSIs*…. QIT08”. They also, all together agreed that the SSI syndromic surveillance is useful in mapping and understanding the source of infection and therefore take appropriate measure.

**Subtheme C: Resource allocation** by Proper planning of strategies that will **Improvement in IPC** in PHFs:

In order to enhance IPC, a comprehensive plan should incorporate a multimodal approach suggested by WHO with essential elements like as system change, education, monitoring, and feedback while cultivating a strong safety culture. A staged strategy is required, beginning with evaluation, followed by action plan development, implementation, impact assessment, and long-term sustainability. This is in keeping with the view of one respondent who said… “*SSI surveillance is useful as it helps in planning correct decision for choosing the countermeasures and planning for the interventions and strategies of it*…QIT09” another one said… “*Its usefulness is on using resources rationally because you plan based on informed decision*…. QIT10”

Data on many facets of the healthcare system and procedures are essential for planning actions to improve interventions like IPC. This information about infection rates including SSI rate, current IPC procedures, the availability of resources, and the background of the particular community and healthcare facility. One respondent said…. *At all levels where healthcare is provided interventions should be planned based on data collection and analysis to obtain the evidence*,,,,,, QIT11”. One of the participants said… “*SSI surveillance is useful because data collection about any infectious disease outbreaks or SSI that occur within the health facility helps to comprehend the nature and how bad it is and carry out focused interventions*…. QIT05”

Resources like PPE and other equipment are crucial for preventing SSIs as one of the IPC measures available. One of the respondents said…. *You know the usefulness of SSI surveillance, PPE and other IPC related equipment are crucial for mitigation of SSIs and other HAIs, by knowing the extent of SSI will facilitate the availability of PPE and other equipment to be available all the time*…. QIT26” Another participant said…. “*It is necessary to understanding the problem of SSI and its extent so as to determine the number of PPE and other supplies needed. Also, SSIs is essential for avoiding and detecting infections following surgery*…. QIT27”. One of the participants had a view that……. “*Infections are tracked and reported, which enables HFs to enhance procedures, evaluate the success of prevention measures, and eventually lessen patient harm by improving IPC*…. QIT28”

### Subtheme D: Giving alerting of what may happen in terms of the burden of HCAIs

One of usefulness of the SSI surveillance can be in terms of IPC procedures that can be greatly enhanced by alarming HCWs on the possible burden of HAIs like SSI. Stressing the negative effects of HCAIs, like longer hospital stays, higher expenses, and even death, can encourage HCWs to follow IPC guidelines. One respondent reported….. “*The severity of this problem is demonstrated by data giving alert of high rates of death, extended hospital stays, rising antibiotic resistance, and significant financial implications to individuals and healthcare systems*….QIT13”. One of the respondents thought that… “*Another usefulness of SSI surveillance is on the data that presents an alarming picture of the incidence of rates of SSI, highlighting the necessity of heightened awareness, prevention initiatives, and continue with strong surveillance systems to lessen the impact of these SSI on our surgical patients*…QIT12”

Furthermore, usefulness of the surveillance include understanding and controlling the total burden of SSI which depends heavily on SSI surveillance. One participant reported that…. “*By monitoring SSI cases, surveillance offers important information about the rates, risk factors, and consequences of SSI, which aids in the creation and assessment of IPC plans…* QIT14*”* These facts were echoed by other respondents, whereby one of them said…. “*The beauty and usefulness of the SSI surveillance is what it does, it is showing what is happening and status of SSI and as well demonstrate the status of IPC in our facility*…QIT15” furthermore another member said…. “*In essence, SSI surveillance is useful because it helps to evaluate ourselves how well IPC strategies are being implemented in a surgical context in our facility. Monitoring SSI rates, spotting possible issues with IPC procedures, and applying the knowledge gained to enhance such procedures are all part of it*…. QIT16”.

The SSI surveillance is useful in monitoring infection rate trends. The respondents said “…. *To do this, the number of SSIs is tracked over time in order to spot trends, evaluate how well preventative measures are working, and maybe spot outbreaks*…. QIT18 Some respondents reported that…. “*Healthcare facilities can identify particular surgical procedures, departments, surgeon or times when SSI rates are higher than anticipated by examining trends. This enables to concentrate the efforts on specific initiatives aimed at lowering infection rates*…QIT17”

### Subtheme E: Improvement of health services quality in the PHFs

Surveillance for SSIs is essential to raising the standard and therefore quality of health services, especially in surgical settings. The participants said…*Surveillance helps locate areas for improvement in IPC procedures by tracking and evaluating post-operative infection rates. This, in turn, improves patient outcomes and lowers healthcare expenses*…. QIT19. Some of the respondents said…. “*In surgical settings, SSI surveillance is useful in facilitating early detection, focused interventions, and ongoing quality improvement, making it a potent instrument for raising the standard or quality of healthcare. Infection rate reduction, better patient outcomes, and resource optimization are all greatly aided by it*…. QIT20” Moreover, SSI surveillance is a useful instrument for assessing how well IPC procedures are working and raising the standard and quality of surgical services. some participants reported that …. “*Moreover, it is useful in a way that it is evaluating the health services*…. QIT21.” And another said “Healthcare *facilities may guarantee that patients receive the safest and best care possible by keeping an eye on SSI rates, determining risk factors, assessing solutions, and enhancing patient outcomes*…. QIT22”

Surveillance of SSIs is Useful for guaranteeing adherence to established Guidelines, protocols and Standard Operating Procedures (SOPs) for IPC in surgical settings and other areas of the health facility. Respondents said “….. By keeping an eye on the incidence of SSIs, medical facilities can spot instances where procedures depart from advised guidelines and implement remedial measures. Some of the participants said…QIT23”. Others said “……*the usefulness of surveillance is that, data can reveal particular practices that are not recommended by the guidelines and protocols, such as poor hand cleanliness, inappropriate surgical clothing, or a failure to give prophylactic antibiotics at the appropriate time*…QIT24” SSI surveillance directly supports the goal of putting patients’ health first. Some respondents reported…. “*Through IPC, we prioritize better patient outcomes, increased overall quality of care and improving overall health services, that is another usefulness of it*……QIT25”

### Second Theme: Ease of Use, Behavioral intention and actual use of syndromic Surveillance of SSI in PHFs

The implementation and adoption of syndromic surveillance for SSIs within PHFs were heavily influenced by the core constructs of the TAM: Perceived Ease of Use (PEOU), Behavioral Intention (BI), and Actual Use (AU). Because PHC settings often face severe resource constraints, understanding these relationships is critical for transitioning from passive reporting to active, pre-diagnostic tracking

### Subtheme A: Provision Data-driven and evidence based on how best to plan for the counter measures

An effective countermeasure plan begins by gathering baseline metrics and historical data to isolate specific root causes. Planners then select interventions that have a proven, documented track record of mitigating those exact risks. An economic appraisal follows to ensure the selected strategies yield a favorable cost-benefit ratio before deployment. Finally, implementation relies on continuous real-time monitoring of performance indicators to iteratively refine the system.

Specifically, SSI surveillance gives patients and HCWs vital information about the causes of post-surgical infections, allowing them to adopt appropriate precautions and treatments. However, for syndromic surveillance to be successful, the entire process must be user-friendly so that HCWs can easily utilize the system. Some respondents reported that… “*it is easy to use of SSI surveillance system educating HCWs to understand risk factors, possible causes, and suitable interventions to reduce the spread of infections and enhance patient outcomes*……QIT26”. Another respondent said…… “*SSI surveillance used to give HCWs insightful input that helps them enhance patient safety and resilience in healthcare environments by lowering infection rates and improving practices* QIT27”

The SSIs is essential for educating HCWS and making them aware about infection risks and prevention techniques, which will eventually enhance patient safety and care. One respondent said…. “*HCWs practitioners can better avoid SSIs and enhance patient outcomes by using SSI surveillance, which offers feedback on performance and adherence to standards* QIT28” The respondents said …… “*Surveillance is used to facilitate communication between different healthcare teams involved in a patient’s perioperative care, ensuring everyone is aware of the infection risks and prevention strategies*……QIT29” another said… “*We use SSI surveillance in determining the magnitude of SSI cases and assessing the results of SSI prevention initiatives*…. QIT31” Also another emphasized that “… *we use the surveillance for figuring out how big the magnitude is, figuring out risk factors, and evaluating how well preventative measures are working*….QIT33”. Aother said “…*we use to determine the scope of the problem, identify areas that require improvement, and assess the results of adopted interventions by gathering and evaluating data on SSIs*…. QIT32”

### Subtheme B: Facilitating and constricting factors for use of syndromic Surveillance of SSI

The use of this system is highly dependent on environmental and institutional conditions. Facilitating factors like strong leadership, adequate resources, and positive peer encouragement significantly drive user adoption. Conversely, constraining factors such as severe time limitations and resource shortages frequently hinder successful implementation. Additionally, shortage of HCWs creates a substantial obstacle to routine integration.

Surveillance of SSIs use is facilitated by the presence of infrastructure, trained HCWs, importance of data for decision. The respondents reported that…. “*Collecting SSI data in our HFs is ease because we have registers, summary forms for monthly and quarterly reporting, that makes our work simple* QIT34. Another respondent said…… “*We were trained by the ministry of health to undertake that obligation; therefore, we are conversant with the entire process*…. QIT35”. Furthermore, one participant reported…… “*We are getting support from higher authorities and fellow HCWs in implementing the syndromic surveillance*…. QIT36”. Another said…. “*We also know surveillance data is valuable tool for benchmarking, allowing hospitals to compare their SSI rates, identify areas for improvement, and ultimately enhance patient safety and care…*. QIT37”. In addition, another participant said……… *benchmarking against peers helps identify best practices and areas where performance can be improved through experience sharing*…. QIT40”.

“SSI surveillance is essential to monitoring and enhancing surgical safety and determines what to improve through evidence. Moreover, SSIs are prevented, patient outcomes are improved, opportunities for improvement are identified, planning to rectify the problem or source and infection rates in various settings are benchmarked……” Some of the respondents suggested that…… “*Surveillance of SSI is used as an essential tool for cultivating a quality-improvement mindset in healthcare, especially in surgical settings. By monitoring and evaluating SSI data, HCWs can find problematic areas, carry out focused interventions, and eventually improve patient outcomes*…. QIT38. Changes in methods and procedures are data-driven insights in a culture of continuous quality improvement that is fostered by Surveillance of SSI. Other respondents perceived that…. “*Patient-centeredness is improved by SSI surveillance. By monitoring SSIs, HCFs can identify areas for surgical care improvement and implement policies that prioritize patient safety and well-being. It is also used to reduce the risk of SSIs and improve patient outcomes, this means optimizing pre-, intra-, and post-operative care*…. QIT39”

Despite the facilitating factor, there were also some constricting factors in the implementing of the syndromic surveillance. The participants said… “*We have limited time to implement and oversee the syndromic surveillance and IPC implementation in general, this is because we are involved in other duties*…. QIT38. Another one added…… “*the shortage of HCWs is the reason that the implementation of syndromic surveillance becomes difficult*……QIT41”.Another respondent said….” *the tools we use are not so friendly that we can’t complete filling leading to incompleteness and incorrectness*……QIT42”. Another reported that…. “Use *of the semi-automated tools (Hard copies of registers and summary forms and ultimately fill in the digital platyform DHIS2) is somehow cumbersome and constricting the effectiveness of the execution of the syndromic syveillance*……QIT43”

### Subtheme C: Frequency and consistency of surveillance reporting

Some participants reported… “*Surveillance of SSI if used consistently, ultimately contribute to better surgical quality and patient safety by tracking and reducing the incidence of SSIs. It also helps achieve other standards by providing data for benchmarking, identifying areas that require improvement, and focusing efforts on quality improvement*…. QIT44”. Others also reported…… “SSI *surveillance when actually used helps lower the risks of SSI and is an essential part of successful IPC programs. By keeping an eye on SSI data, HCWs evaluate the success of solutions put in place, and eventually improve patient safety* QIT45”. Another said…. *SSI surveillance when used consistently makes HCWs to be dedicated to patient safety and high-quality care and foster trust from patients*…… QIT46. Some respondents said……. “*HCWs mitigate the risk of infections by actively tracking and evaluating SSI rates. This enhances patient outcomes and boost trust in the healthcare system* QIT47”

### Subtheme D: Identification of gaps related to implementation and compliance of IPC by HCWs

Surveillance of SSI is an essential technique for detecting gaps in HCWs’ compliance and application of IPC. Monitoring the frequency and features of SSIs enables surveillance activities to identify areas with inadequate or irregularly implemented IPC procedures, enabling focused interventions to enhance infection control procedures and lower SSI rates. One respondent reported…… “*SSI surveillance essentially used as a feedback system, giving us important information about gaps in IPC measures and empowering us to take preventative measures to improve IPC and surgical results*…… QIT48. “Some other respondents reported… “*Hospital safety is greatly enhanced by SSI surveillance, which helps in the detection, avoidance, and management of SSIs. Our hospitals can lower the incidence of SSIs and enhance patient outcomes by implementing focused interventions after tracking and evaluating infection rates* QIT49”

### Subtheme E: Provision evidence based on how best to plan for the counter measures

Surveillance SSI is used to provide essential data for developing and implementing effective strategies to prevent and control SSIs. Surveillance systems systematically monitor and analyze data on SSIs to identify trends, risk factors, and areas for improvement, hence assisting healthcare facilities in tailoring their actions. This evidence-based approach is essential to enhance patient safety and reduce the burden of SSIs. Some of the participants said… “*The surveillance is used as one of a key instrument for strengthening hospital resilience using evidence, which lowers the incidence of SSIs and improves IPC procedures. Hospitals can bolster their defenses against HAIs by tracking SSI rates and determining areas that require improvement*…. QIT34”. Another respondent emphasized by noting…. “*SSI surveillance used to provide accurate data for effective infection rate benchmarking*…. QIT21”

Enhancing patient outcomes, assessing preventative initiatives, and identifying SSI instances are all made possible via surveillance…. Some other participants said… “*SSI surveillance is used as a crucial component of safe surgery practices since it helps track and reduce the incidence of SSIs and enhances patient outcomes. By tracking SSI rates, hospitals can identify areas with evidence for improvement, planning to rectify the problems, implement targeted treatments, and assess the effectiveness of their IPC strategies* QIT4”

### Subtheme F: Provision of knowledge on the source of infection to the patients who undergo surgery

Surveillance of SSI data is used to identify problems, start remedial actions, and ultimately reduce the incidence of SSIs. By tracking SSI rates, healthcare facilities can assess their performance, determining what to do in order to improve patient outcomes by evidence. Some respondents reported… “*SSI surveillance is mostly used for determining the etiology in terms of cause and source of infections as well as number of those who get infected. By carefully tracking and analyzing SSI instances, healthcare facilities can locate potential contamination sources, such as specific surgical instruments, environmental factors, or healthcare staff behaviors*…… QIT 19. “Also, some participants said…. “*SSI surveillance is used as more than just collecting data, is a dynamic process that improves patient outcomes by educating, guiding, and enhancing IPC practices*……QIT22”. Some of the respondents claimed that…… “*SSI surveillance is used to make the HCWs be aware at every time when they evaluate the success and gaps of their current preventative initiatives by tracking SSI rates over time. It is necessary to reassess and modify the strategy, possibly introducing new rules or training, if rates are not declining or are even rising. Other respondents said*……QIT2. “It is used to monitor infections that develop after surgery, which is essential to determining their prevalence and trends.

## Discussion

### Usefulness of implementation of syndromic Surveillance of SSI

Syndromic surveillance for SSI is useful for early detection of potential outbreaks, tracking disease trends, and informing public health responses(19,20). It helps streamline collaboration between public health and healthcare groups, improve understanding of health inequities, and potentially lead to more effective policymaking and resource allocation. Syndromic surveillance provides a way to identify potential outbreaks of SSI before they are confirmed through traditional laboratory-based surveillance, allowing for a quicker and more effective response(21). It can augment existing surveillance systems, like event-based surveillance using alerts and rumors, by providing additional data and insights. By monitoring trends in symptoms associated with SSI, syndromic surveillance can help public health officials understand the scope and impact of these infections in a community. The respondents’ opinion was in keeping with the review that reported, the SSI surveillance framework provides the systematic steps of documentation and reporting of the SSIs occurrences from the surgical patients. In the process of documentation, potential outbreaks, trends can be established (22)

The system can facilitate better communication and coordination between public health agencies and healthcare providers, leading to a more efficient responses. Study by KD Mandl, also concluded that Syndromic surveillance technology is useful not only for bioterrorism event detection, but also for general public health, clinical medicine and for quality of health improvement(20). Early detection through syndromic surveillance can help optimize resource allocation by identifying areas with high SSI rates and targeting interventions more effectively. Early detection and intervention can potentially reduce the overall cost of treating SSI by preventing more severe infections and hospitalizations(23,24). Syndromic surveillance systems can be adapted to monitor a wide range of health events, including not just SSI but also other infectious diseases and public health concerns. The great sensitivity of syndromic surveillance helps offset the low sensitivity of laboratory-based surveillance techniques(25). Using syndromic surveillance in conjunction with laboratory-based surveillance techniques will significantly help determine the actual burden of HAIs with restricted resources. Data-driven decision-making and resource allocation are also made easier by its adaptability to various healthcare settings and connection with current health information systems(23,26,27). To detect potential outbreaks early, such a system would supplement the event-based surveillance method, which relies on rumors and alarms. The expense and burden of the HAIs’ consequences may be decreased if priority HCAIs, such as surgical site infections, were monitored in a more efficient and focused manner. With standardized data collection, healthcare worker training, real-time reporting systems, stakeholder participation, and ongoing monitoring and evaluation, this strategy might be extended into national HCAI surveillance and provide early detection capabilities. One promising tactic for preventing HCAIs in LMICs like Tanzania is syndromic surveillance. It improves patient safety, saves resources, and offers early warning capabilities. Strategic interventions, stakeholder cooperation, and continual monitoring and assessment are necessary for effective implementation in order to guarantee long-term efficacy in HCAI detection and response(26).

Early outbreak identification, tracking the magnitude, spread, and pace of epidemics, tracking disease patterns, and ensuring that an outbreak has not occurred are all possible using syndromic monitoring.

Syndromic surveillance systems aim to leverage current health data in real time to give those in charge of looking into and monitoring possible outbreaks quick analysis and feedback(23,27,28). For certain diseases, the optimum data sources for outbreak surveillance and the ideal symptom criteria for ongoing monitoring have not yet been identified. The goal is to find widely applicable signal detection techniques and response protocols that would optimize detection while conserving limited resources. Early outbreak identification, tracking the magnitude, spread, and pace of epidemics, tracking disease patterns, and ensuring that an outbreak has not occurred are all possible using syndromic monitoring. Syndromic surveillance systems aim to leverage current health data in real time to give those in charge of looking into and monitoring possible outbreaks quick analysis and feedback. For certain diseases, the optimum data sources for outbreak surveillance and the ideal symptom criteria for ongoing monitoring have not yet been identified. The goal is to find widely applicable signal detection techniques and response protocols that would optimize detection while conserving limited resources (27).

### Ease of Use, Behavioral intention and actual use of syndromic Surveillance of SSI in PHFs

The respondents perceived the use of syndromic surveillance of SSI to be easy and important. They all reported syndromic surveillance systems monitor SSI symptoms almost instantly and enable prompt detection of possible outbreaks or infection clusters. It offers a heads-up for public health response by detecting possible SSIs before test confirmation. It facilitates the monitoring of SSI patterns over time, enabling the evaluation of IPC strategies’ efficacy(19,29). By supplementing conventional techniques like lab confirmation and case reporting, syndromic monitoring can provide a more complete picture of the burden of SSI. Syndromic and conventional surveillance systems are frequently combined, utilizing the same infrastructure to collect data on a range of public health incidents. Rapid intervention following early SSI discovery may lower morbidity and mortality. These opinions from respondents match with the opinion of Hughes, H.E., et al from the systematic review which they concluded that Syndromic surveillance provides a valuable tool for the identification and monitoring of trends in severe illness. Technological advances, particularly in the emergency care patient record, have enabled the evolution of trends over time. syndromic reporting has become closer to ‘real-time’, with automated, secure electronic extraction and analysis possible on a daily, or more frequent basis (30).

Better knowledge of the SSI status in their PHFs can help facility management plan responses and allocate resources more efficiently. Syndromic monitoring can lessen the strain on conventional surveillance techniques by offering an early warning system, particularly in times of public health emergency(31). Syndromic surveillance provides rapid, thorough information on possible infections, making it a useful method for SSI surveillance in PHFs. Despite its drawbacks, when used properly and morally, it can be a very useful tool for early detection, surveillance, and outbreak response(32–34). The results also suggest that the syndromic surveillance is helping to address gaps for policymakers, decision-makers, program implementers, and advocates in the IPC stakeholders for accessible, practical, and use of information to support the design and implementation of evidence-based IPC policies and programs. In the global health space, this finding reminds all stakeholders to take steps and make sure all health facilities that conducts surgeries to have any form surveillance so as to trace SSI and develop interventions which will help to address those gaps based on evidence.

### Quality Control

1. Pre-testing the guiding questions and checking for their completeness was done daily.
2. Translation and back translation of questionnaires was done.
3. Back up of the data and filled questionnaires was stored in a safe place under lock and Key.
4. Meetings with research assistants were done to sort out data collection problems.
5. Interviews were conducted by a trained set of data collectors who were fluent in the local language, formally trained in a quantitative and qualitative research workshop, and who were having previous experience in the qualitative data collection process. They will be trained on the study methodology by the investigators.

## Conclusion

The SSIs in Tanzanian PHFs involves the systematic tracking of clinical signs such as wound redness, purulent discharge, and fever rather than relying on delayed microbiological laboratory testing. This approach is highly practical in resource-limited settings where advanced diagnostic equipment is often unavailable or inaccessible. By empowering HCWs workers to rapidly document these observable symptoms using standardized reporting tools, health centers can bridge critical diagnostic gaps and identify potential infections shortly after procedures.

The primary usefulness of this proactive monitoring lies in its ability to facilitate early detection and prompt management of SSI, which significantly reduces patient morbidity, readmission rates, and healthcare costs. In Tanzania, where rural geography and limited transportation can make patient follow up challenging, syndromic data enables district health teams to spot localized infection clusters early, optimize antibiotic stewardship, and improve surgical safety protocols. Ultimately, this continuous monitoring strengthens the broader health system.

## Data availability

The corresponding author can provide the data that is supporting the study’s conclusions upon request at any time.

## Author Contributions

JCH: Conceptualization, Methodology, Writing of Original Draft, Funding Acquisition data collection, analysis and report writing

## Ethics statement

The research plan was approved by the Tanzanian Ministry of Health (reference number PA 131/142/01/412) and the President’s’ Office - Regional Administrative and Local Government (reference number AH. 161/323/01/50). Also, the National Institutional for Medical Research granted ethical clearance (reference number NIMR/HQ/R.8a/Vol. IX/4687). In addition, study identification numbers were used instead of using the participants’ names to maintain their anonymity. The gathered data were safely preserved on a password-protected computer and in physical copy in a lockable drawer.

## Data Availability

Data can be accessed upon request

## Acknowledgement

I would like to thank the Almighty God for giving me strength, guidance, and courage throughout the period of this research development. Secondly. I would like to thank my employer, the Ministry of Health of the United Republic of Tanzania for allowing me to conduct this research.

